# Is ultrasound imaging effective in teaching anatomy in undergraduate medicine? A systematic review protocol

**DOI:** 10.1101/2022.11.02.22281829

**Authors:** Eloise Powell, Lucy Hammond, Frederick Marlowe

**Affiliations:** University of Warwick

## Abstract

Anatomy knowledge is a foundation of learning medicine and is traditionally taught to students using cadaveric dissection. In the last few decades, a variety of adjuncts to teaching anatomy have been developed including plastic models, plastinated human specimens, living anatomy (surface) and radiological images to aid learning. Portable ultrasound (US) has become a useful learning tool that is safe and non-invasive allowing for visualisation of organs and associated structures. The role of ultrasound has been widely discussed in the literature with some institutions integrating it into the medical curriculum.

This protocol describes a planned study that aims to evaluate the effectiveness of ultrasound in teaching anatomy to medical students by systematically reviewing the existing literature available in the public domain. Data gathered by these studies can be extracted and analysed to provide evidence of the effectiveness of ultrasound in medical education. The outcome will potentially support medical educators in integrating ultrasound aided learning into the curriculum with the aim of improving students understanding of anatomy. A secondary outcome may be basic understanding and competency in ultrasound.

## Background

In medical curricula, X-ray and CT imaging is often used to supplement traditional anatomy teaching (Lufler *et al*., 2010). Over the last decade or so some medical school programmes have been making efforts to integrate ultrasound teaching into their curriculum. However, several barriers to ultrasound use have been described; perceived effectiveness of ultrasound education and resources for ultrasound technology being examples.

Research by Swamy & Searle (2012) demonstrated that introduction of adjunct ultrasound teaching with line diagrams significantly improved students’ perceptions of their understanding of anatomy. Dreher et al (2014) found that this increased student perception of confidence in using ultrasound and understanding of anatomy was congruous with performance on practical examinations however the researchers did not report on the statistical significant of this and so it is unknown if the training is successful of demonstrating learning had occurred. These results were found again by DesJardin et al (2017) who assessed the validity of a new ultrasound course for first year medical students. Students perceived ultrasound to be useful in learning anatomy, a view supported by improved performance on the post-course test, however these results were not statistically significant.

Dinh et al (2015) successfully implemented an ultrasound curriculum demonstrating significant improvement in objective structed clinical examination (OSCE) performance compared to historical data. These finding were supported by a recent study demonstrating a statistically significant improvement in test scores in the ultrasound group following a simulation versus didactic teaching session (Shah *et al*., 2019). Alexander et al (2021) found that ultrasound was a useful educational tool when teaching anatomy to non-medical students as well.

Kondrashov et al (2015) found that students who undertook a clinical ultrasound elective course had significant improvement in their post-test anatomy exam score in comparison to their pre-test. Further discussion identified that there were learning areas that were more successful for student knowledge with scores for the neck and eyes showing no significant improvement. However, Griksaitis et al (2012) demonstrated that using portable ultrasound to demonstrate cardiac anatomy was equally as beneficial, although not better, to traditional cadaveric teaching using pre- and post-test scores following the teaching intervention.

One study looked at the feasibility of integrating ultrasound teaching on cognitive load on students (Jamniczky *et al*., 2017). The learners exposed to the ultrasound teaching performed significantly better compared to the historical cohorts without negatively impacting learning. There is substantial evidence to suggest that students are supportive of the integration of ultrasound into the medical curriculum in a number of areas such as physiology, anatomy, clinical skills and procedures. A recent scoping review found widespread support for the integration of ultrasound into undergraduate medical education. They surmised that ultrasound had the benefit of showing *‘‘living anatomy’ through dynamic representations of anatomic structures and their relationships ‘*(Tarique *et al*., 2018). The researchers reported that students regarded the incorporation of ultrasound teaching favourably and that they felt their knowledge and understanding of anatomy was increased as well as confidence identifying structures. However, they did not review the role of ultrasound use in teaching anatomy in depth or compare the ultrasound exposure to existing anatomy teaching. The researchers did not comment on their critical appraisal of the available literature and their discussion was focused around the positive student perception and no other measurable outcomes.

Undertaking this systematic review has the potential of supporting integration of US imaging into the medical curriculum, improving students understanding of anatomy and developing basic competency using ultrasound (Frank, 2010).

There is a large body of existing research regarding the integration of ultrasound into the medical curriculum and whether it benefits student learning. Studies measure effectiveness of ultrasound teaching using different educational outcomes. To the researcher’s knowledge there is not a systematic review of the literature relating to effective use of ultrasound in teaching anatomy to medical students at present. Therefore, the aim of this research is to systematically review and evaluate if ultrasound is effective in anatomy teaching in undergraduate medicine. Effectiveness will be measured using Kirkpatrick’s 4-level model of evaluation. This can inform future teaching practice and potentially integration of ultrasound into the medical curriculum. A secondary outcome from this study may be improved basic understanding of ultrasound and competency in using ultrasound.

### Project approach, methods and analysis

#### Study design and data collection

This study will be a systematic literature review +/- meta-analysis +/- meta-synthesis of the available data covering the effectiveness of using ultrasound in teaching anatomy and will be undertaken over a period of 8 months.

- Population – undergraduate medical students
- Exposure – ultrasound teaching of anatomy
- (Comparison) – existing anatomy teaching/traditional anatomy teaching - this may be lecture-based with cadaveric dissection, plastinated pro-sections, medical imaging etc.
- Outcome – effectiveness of the teaching, with outcomes categorised by Kirkpatrick’s 4-level model of evaluation (Kirkpatrick, 1996) (see below). Note that outcome will be reviewed when screening the literature obtained through the database search.

| Kirkpatrick Level | Outcome measurement |
| --- | --- |
| 1 – Reaction | Increased learner satisfaction or confidence of anatomy knowledge |
| 2 – Learning | Increased knowledge of anatomy |
| 3 – Behaviour | Influence of teaching on application of learning (ability to apply anatomical knowledge to other areas of medicine) |
| 4 – Results | Improvements in clinical practice and benefits to patients |

Studies available for review will be identified through several databases: Embase, Medline, Web of Science and Education Research Complete.

Search terms will include key words to capture all relevant studies:

- Ultrasound or US or point-of-care ultrasound or ultrasound imaging or US imaging or portable ultrasound or portable ultrasound imaging or focused portable ultrasound imagine or sonography or sonograph or ultrasonic therapy or ultrasonography or ultrasonography or echography or echograph or handheld ultrasound device or insonation
- And
- Anatomy or anatomical
- And
- Teaching education or learning or teaching or curriculum

If many studies are returned from the initial search (n>60) students will be included as a search term. Variations may include undergraduate medical student or education or undergraduate education or undergraduate medical education or medical curriculum/curricula or undergraduate medical curricula or clerkship or pre-clerkship or pre-clinical or clinical.

These search terms may yield studies that also apply ultrasound to other themes of medical education, e.g., teaching clinical skills or procedures. Boolean operators will allow us to include these papers in the initial search and then review by the researchers can identify their suitability for inclusion/exclusion.

#### Inclusion criteria

Studies that include ultrasound imaging and real-time ultrasound teaching, studies that use qualitative or quantitative methodologies, are global, are peer-reviewed published articles, are specific to medical students, written in English language and are empirical research studies and primary research studies. Studies whose outcomes align with Kirkpatrick’s framework will also be included.

#### Exclusion criteria

Studies involving postgraduate teaching/training, studies where the participants are not medical students, studies which include graduate entry medics who may have previous experience of ultrasound, studies not dedicated specifically to an anatomy curriculum, studies awaiting publication, studies whose data is not extractable, studies published in a non-English language and non-research studies including case reports and educational commentaries, opinions pieces, literature reviews, any study that has outcomes which do not fall into the Kirkpatrick levels defined above, will be excluded.

#### Search Strategy

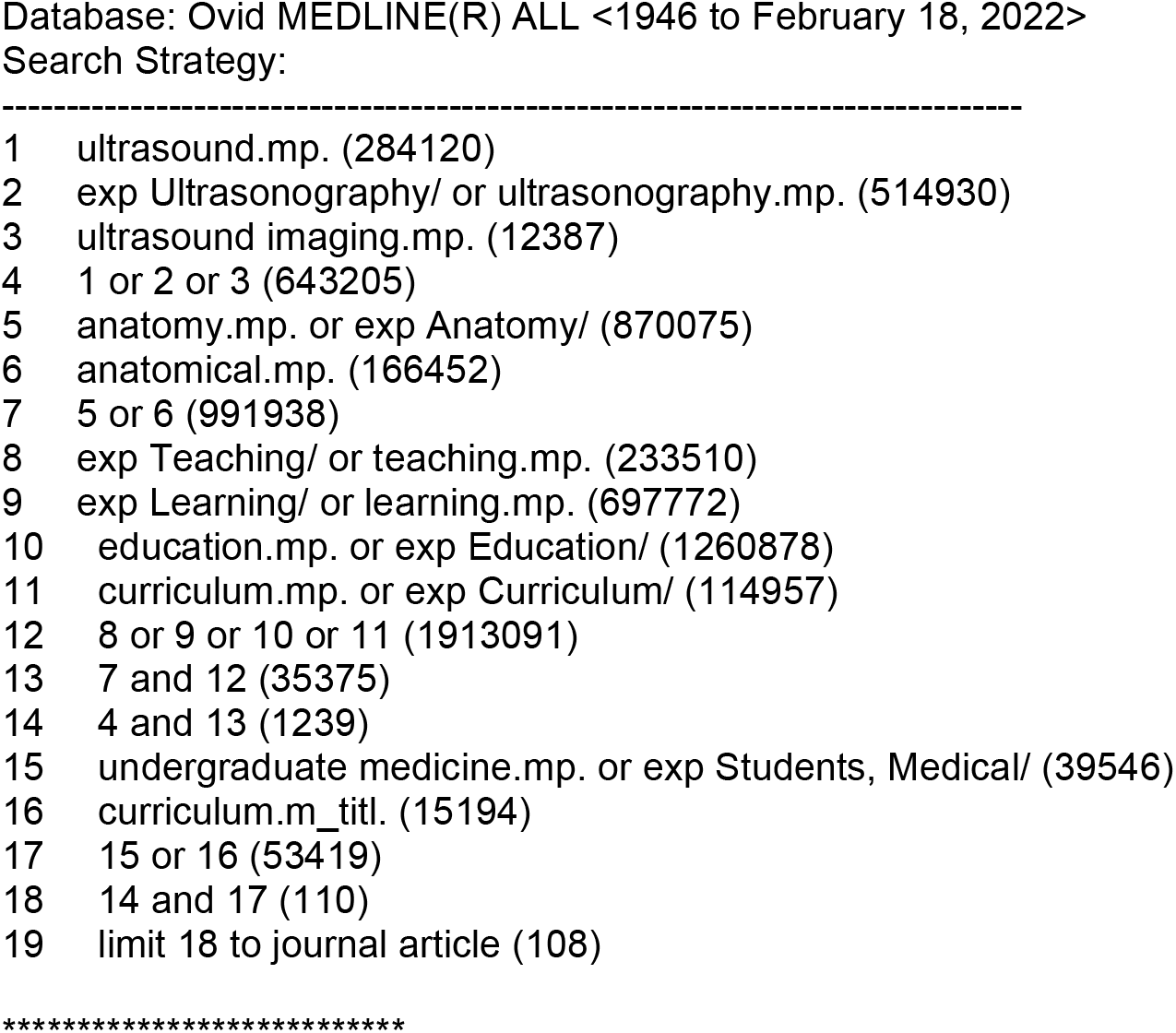

#### Screening and selection

All articles identified from the preliminary search will be reviewed for eligibility by the researcher and a second independent reviewer. The project supervisor will act as the third reviewer if there is discordance in opinion for inclusion.

Studies identified may be eligible for qualitative synthesis and quantitative analysis. These studies will be included if they can demonstrate outcomes related to Kirkpatrick’s 4-level model of evaluation (identified literature may fall into multiple categories).

Once the database searches have been performed, citations will be exported into Endnote and duplicated studies will be removed. In line with the PRISMA-P framework the citations will then be imported into Rayyan for primary screening involving title and abstract screening (Shamseer et al., 2015). Following this the researchers will screen the full texts for inclusion/exclusion using Covidence.

Quality appraisal of relevant content and methodology in the identified studies will be performed using JBI critical appraisal checklist to assess for validity and bias (Joanna Briggs Institute, 2017).

#### Analysis and synthesis

Analysis will be a narrative synthesis that may include meta-analysis and/or meta-synthesis. This will be determined once the database searches, screening and full-text review has been has been performed and it is known whether meta-analysis and/or meta-synthesis is suitable. Data will be extracted using the Covidence web-tool. Categories with a few examples have been developed to guide the data extraction process:

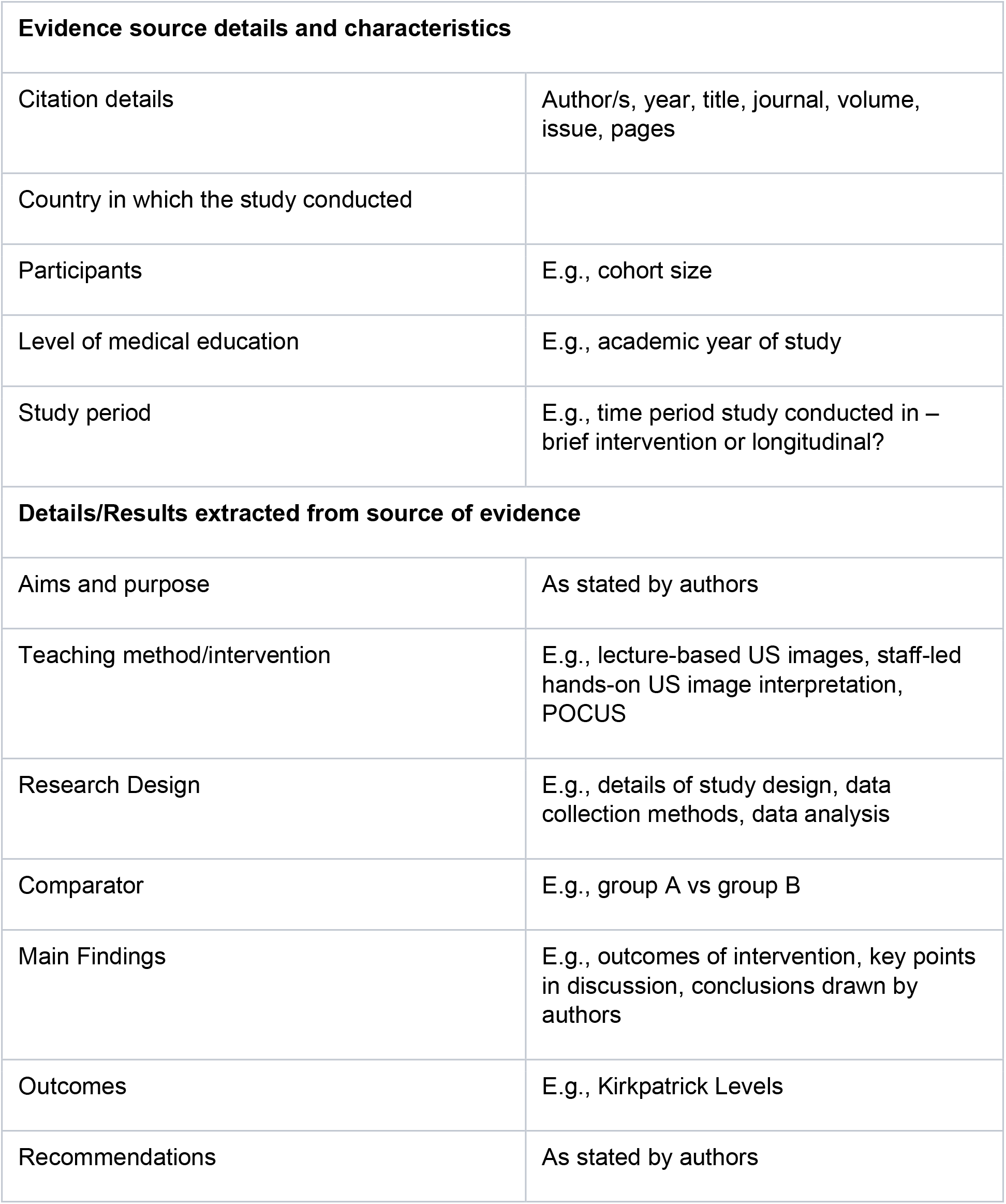

Kirkpatrick’s 4-level model of evaluation will be used to structure the discussion of the findings of the systematic literature review.

### Data collection, use & storage

Data collected will be by a systematic search in the databases listed above. As no new data will be generated during this study there are no concerns with personally identifiable data or need for data protection governance. Data will be stored on the researchers’ computer and will be accessed by the primary researcher.

### Risks & ethical considerations

No formal ethical approval is required for this project as the data collected is available in the public domain. Only published material will be used in this study.

## Dissemination

The results of the study will be submitted to either an educational or radiology related journal. Appropriate education journals may include Medical Education, The Clinical Teacher, Medical Teacher, BMC Medical Education. Appropriate radiology journals may include British Journal of Radiology, Imaging, European Journal of Imaging, Clinical Radiology journal, Academic Radiology.

The study will also be submitted to potential educational or relevant radiological conferences for presentation or poster presentation.

## Supporting information

PRISMA-P 2015 Checklist

## Data Availability

All data produced in the present study are available upon reasonable request to the authors

## Changes to the Protocol

The author does not anticipate any change to the protocol at this stage. Any changes will be explained and justified in the final written article.

